# Delivery of CPAP respiratory support for COVID-19 using repurposed technologies

**DOI:** 10.1101/2020.04.06.20055665

**Authors:** P. Culmer, A. Keeling, C. Osnes, W. Davis Birch, D. Jones, I. Waters, R. Hetherington, J. Parmar, T. Lawton, S. Ashton, M. Latham, S. Murdoch, D. Brettle, N. Kapur

**Affiliations:** University of Leeds, Leeds, UK; Leeds Teaching Hospitals Trust, Leeds, UK; Bradford Teaching Hospitals NHS Foundation Trust, Bradford, UK

## Abstract

The COVID-19 pandemic has placed a dramatic increase in demand on healthcare providers to provide respiratory support for patients with moderate to severe symptoms. In conjunction, the pandemic has challenged existing supply-chains to meet demands for medical equipment and resources. In response to these challenges, we report our work to repurpose two existing non-invasive ventilation (NIV) systems to provide solutions for the delivery of oxygen-enriched CPAP ventilation which are inherently resource and oxygen-efficient. We consider adaptation of CPAP systems typically used for sleep apnoea, together with a new Venturi-valve design which can be readily produced through 3D printing. Our aim in both cases was to support Positive end-expiratory pressure (PEEP) of ≥10cmH_2_O while achieving ≥40% FiO2. This supports a crucial part in the patient pathway for COVID-19 treatment, helping to provide early respiratory support prior to invasive ventilation options in the ICU.

---

The COVID-19 pandemic, caused by severe acute respiratory syndrome coronavirus 2 (SARS-CoV-2), has placed a rapid increase in demand on healthcare providers (HCPs) to provide respiratory support for patients with moderate to severe symptoms [1]. HCPs do not have sufficient ventilator provision to meet this surge in demand. Emerging clinical reports indicate that Continuous Positive Airway Pressure (CPAP) non-invasive ventilation can help patients with moderate symptoms to avoid the need for invasive ventilation in intensive care [2-3], a change to the early impression that early intubation was indicated. Regulatory authorities such as the UK MHRA and US FDA have produced guidance to support rapid development, manufacture and approval of new ventilation systems which can be produced at scale [4, 5]. However, the strains placed by the COVID-19 pandemic on international supply-chains may exceed manufacturing capabilities. Similarly, higher patient numbers in hospitals place increased burden on hospital resources, in particular the provision of medical oxygen crucial for ventilation has faced restrictions to avoid overloading hospital systems and pipework [6]. The oxygen supply issue has been compounded by devices which use oxygen both as a driving gas to power their mechanisms, and directly to oxygenate the patient.

In response to these challenges, we report our work to repurpose two existing non-invasive ventilation (NIV) systems to provide solutions for the delivery of oxygen-enriched CPAP ventilation which are inherently resource and oxygen-efficient. We consider adaptation of CPAP systems typically used for sleep apnoea, together with a new Venturi-valve design which can be readily produced through 3D printing. Our aim in both cases was to support Positive end-expiratory pressure (PEEP) of ≥10cmH_2_O while achieving ≥40% FiO2. This supports a crucial part in the patient pathway for COVID-19 treatment, helping to provide early respiratory support prior to invasive ventilation options in the ICU.

## CPAP machine with O_2_ entrainment

NIV and sleep apnoea (CPAP) machines are widely available to HCPs and provide the potential to deliver therapeutic benefit through enrichment of the air supply. The Nippy 3+ (BREAS Medical) was selected due to its robustness and widespread availability in the locality. In CPAP mode, the system generates 3-20 cmH_2_0 (0.3-2.0kPa) using an internal centrifugal fan to pressurise atmospheric air. This offers the possibility to entrain O_2_ either at the system’s low pressure air inlet, or in the pressurised air-stream near the ventilation mask as shown in Figure 1.

**Figure 1.**
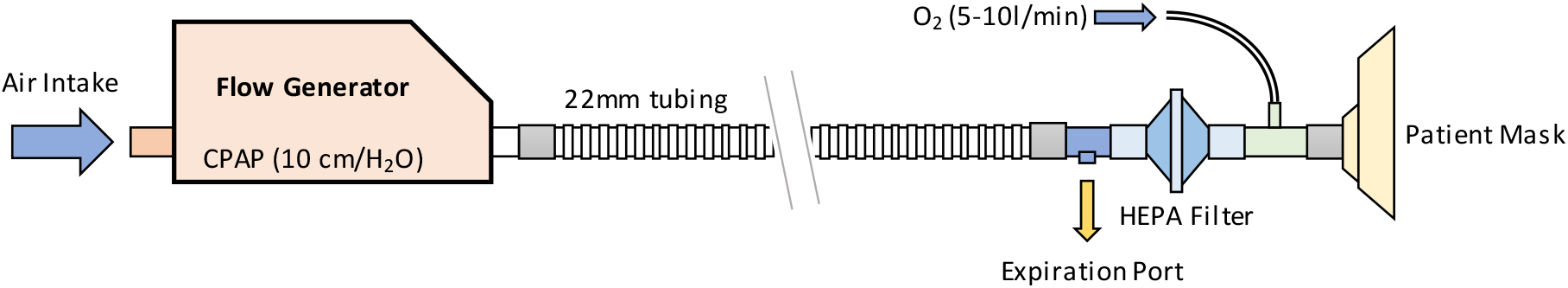
Provision of oxygen-enriched CPAP ventilation using a NIV system

We modified a standard Nippy 3+ system to allow entrainment of oxygen at the intake port and evaluated the efficacy of these two approaches. The system was configured to operate at a pressure of 10 cm H_2_O with 5L/min oxygen flow, combined with a model lung operating at 30 bpm with tidal volume of 0.28L. An oxygen meter was used to measure the effective FiO2 inhaled by the patient.

We have also tested the same approach using the SleepCube (DeVilbiss), an unmodified CPAP machine designed for home use in patients with sleep apnoea. The manufacturer recommends supplemental oxygen be entrained either into the mask, or via an entrainment port at the machine end of the tubing.

## L-Vent: Leeds Venturi valve system for CPAP

Whilst the CPAP machines described above acts as both a flow generator and a pressure regulator, a more traditional circuit uses a flow generator and a separate pressure-controlling valve. The flow generator can be purely mechanical without moving parts; the Venturi effect occurring within a carefully designed valve allows a high pressure oxygen source to entrain and mix air into the breathing circuit. Whilst mechanically simple, these devices can be inefficient in their use of oxygen unless carefully designed. Venturi systems are commonly used in hospitals to provide controlled oxygen therapy without CPAP pressure, however a valve can be designed to cope with both the pressures and flows required for CPAP therapy [7]. While commercial Venturi systems exist, their availability has been limited due to supply-chain challenges during the pandemic. In this resource-limited context, 3D printing provides an ideal alternative means to produce Venturi systems in an agile and timely manner.

For delivery of CPAP for treatment of COVID-19, the design of the Venturi valve must be such that it supports a high flow whilst maintaining a downstream pressure of 10cmH_2_O. This flow rate supports the patients breathing without significantly lowering the pressure. The relative flowrates control the FiO_2_ with 10L/min O_2_ flow and an entrained airflow of 31.4L achieves 40% FiO_2_. Careful design of the valve is required to ensure the pressure requirements are met with previous computational and experimental studies used to guide the design [8]. The L-Vent design was achieved through iterative development in which candidate valves were evaluated for ease of 3D printing and clinical performance in a simple CPAP breathing circuit, as shown in Figure 2. Further information on the L-Vent design and printing process is provided in Supplementary Materials.

**Figure 2.**
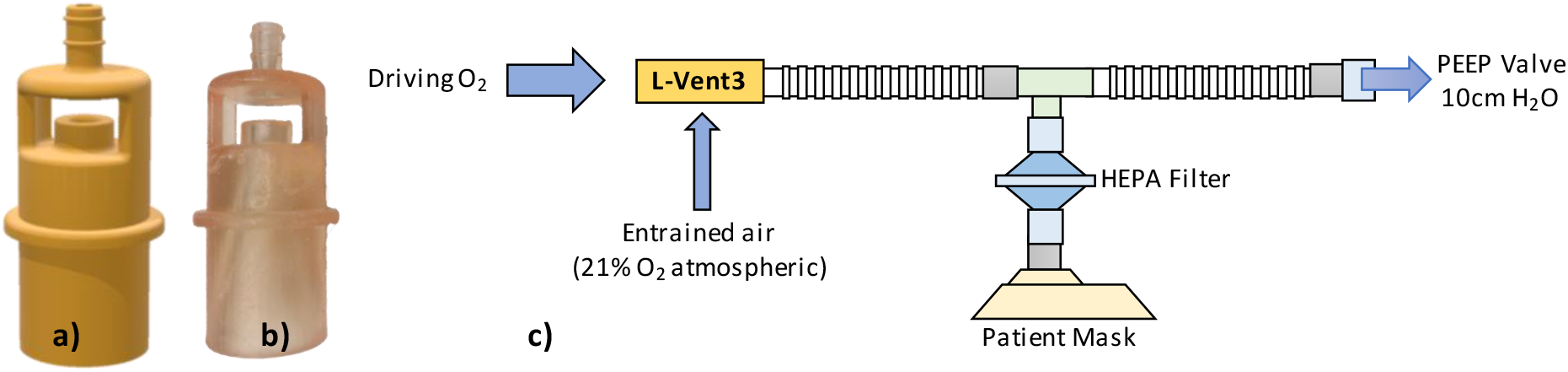
The Leeds-Venturi valve; a) the L-Vent CAD model, b) a 3D printed L-Vent valve, c) L-Vent in a CPAP breathing circuit

## Evaluation

We evaluated both approaches of CPAP delivery to assess their relative merits with respect to resource efficiency and clinical performance.

For the CPAP machines, our results showed that entrainment of oxygen at the low pressure intake brought only modest increases in FiO_2_ (ca. 35%) in comparison to entrainment directly into the circuit which achieved significantly higher FiO_2_ (ca. 50%), with higher values found when entrainment was closer to the mask. The difference in performance can be attributed to the single-arm breathing circuit in which air-flow is reversed during the expiration phase and part of the air column is vented to atmosphere. This results in losses of oxygen when entraining at the air intake, but entrainment near the patient benefits from the pressurised air-column created between the patient and HEPA filter, which acts as a ‘buffer’ to preserve oxygen and avoid losses. Although this testing model precludes gas exchange, it is representative of the relative performance of these two configurations and suggests that relatively low flow-rates of oxygen can be used to obtain therapeutic FiO_2_ levels and that these can be readily altered by altering the oxygen flow-rate.

The L-Vent valve was evaluated using four models 3D printing using a commercially available SLA printer (Form 3, FormLabs) with medically approved dental resin (Dental Model V2, FormLabs). Full information is provided in the Supplementary Materials. Each valve was tested with a driving flow of 10L/min O_2_ in open and closed circuits, representing maximum and minimum flow-rates respectively. Overall, L-Vent achieved 37% FiO_2_ at 50L/min (open) and 44.5% FiO_2_ at 34L/min (closed) and generating sufficient pressure to activate the PEEP valve.

The ultimate FiO_2_ delivered by either system varies with respiratory function and is not explicitly controlled. Thus, these systems require external monitoring by a suitably qualified healthcare professional based upon the patient’s SpO2 level and vital signs in accordance with best practice (e.g. UK MHRA guidance).

## CONCLUSIONS

CPAP ventilation systems provide an important treatment option for COVID-19 patients, particularly in early stages use to deliver oxygen-enriched air to stabilise patients until they can be escalated or de-escalated. To deliver this for the high patient numbers associated with the COVID-19 pandemic, healthcare providers require resource efficient solutions. We have shown that this can be achieved both through the repurposing of NIV ventilation systems and using a novel 3D-printed Venturi system with a standard PEEP-valve breathing circuit. These solutions bring different merits in clinical performance and efficiency, but provide resource-limited healthcare providers with flexible treatment pathways that can be rapidly deployed to reduce the burden on ICU during the COVID-19 pandemic.

## Data Availability

Further data is available from the research team on request.

## Supplementary Materials

- Manufacturing Methods + Associated CAD Files (L-Vent CAD.zip)
- Clinical Usage (L-Vent Usage.pdf)

## Acknowledgements

We would like to thank our multidisciplinary team; Graham Brown, Sam Flint, Kevin Meloy, Mick China, James Naylor, Hardy Boocock, Cath Noakes and the numerous healthcare professionals at Leeds and Bradford Teaching Hospitals (UK) who have helped make this research possible.

## Notes

### Competing Interest Statement

The authors have declared no competing interest.

### Funding Statement

The research is supported by the National Institute for Health Research (NIHR) infrastructure at Leeds. The views expressed are those of the authors and not necessarily those of the NHS, the NIHR or the Department of Health and Social Care.
The authors would like to thank the NIHR MedTech and In Vitro diagnostics Co-operatives (MICs).

## References

1. Xie et al, Critical care crisis and some recommendations during the COVID-19 epidemic in China, Intensive Care Medicine, 2 March 2020 (https://doi.org/10.1007/s00134-020-05979-7)

2. Guidance for the role and use of non-invasive respiratory support in adult patients with coronavirus (confirmed or suspected), NHS England and Improvement, 6 April 2020 (v3)

3. Managing the respiratory care of patients with COVID-19, Italian Thoracic Society (AIPO) and Italian Respiratory Society (SIP), 23 March 2020

4. Life saving breathing aid developed to keep COVID-19 patients out of intensive care, Online Article (https://www.nihr.ac.uk/news/life-saving-breathing-aid-developed-to-keep-covid-19-patients-out-of-intensive-care/24542), Accessed March 30, 2020

5. Galbiati et al. Mechanical Ventilator Milano (MVM): A Novel Mechanical Ventilator Designed for Mass Scale Production in Response to the COVID-19 Pandemics, 31 Mar 2020, arXiv:2003.10405

6. UK MHRA Alert: NHSE/I-2020/001, Accessed 14/04/2020. [www.cas.mhra.gov.uk/ViewandAcknowledgment/ViewAlert.aspx?AlertID=103013]

7. Brusasco et al, CPAP Devices for Emergency Prehospital Use: A Bench Study, Respiratory Care December 2015, 60 (12) 1777–1785; DOI: https://doi.org/10.4187/respcare.04134

8. Eves et al, 2012. Design optimization of supersonic jet pumps using high fidelity flow analysis. Structural and Multidisciplinary Optimization, 45(5), pp.739–745.

